# Analytical Accuracy and Clinical Agreement of a Novel Internet of Things and AI-based Point-of-Care Testing Laboratory

**DOI:** 10.1101/2021.10.29.21264864

**Authors:** Lucca Centa Malucelli, Gabriele Luise Neves Alves, Claucio Antonio Rank Filho, Rafaela Fortes Correa, Vanessa Hintz Albano, Anita Leme da Rocha Saldanha, Tereza Bellincanta Fakhouri, Carolina Melchioretto dos Santos, Matheus Gonçalves Severo, Victor Henrique Alves Ribeiro, Bernardo Montesanti Machado de Almeida, Caio Corsi Klosovski, Tania Leme da Rocha Martinez, Marileia Scartezini, Marcus Vinícius Mazega Figueredo

## Abstract

Point-of-care testing (POCT) offers several advantages over conventional laboratory testing. Nonetheless, a faster turnaround time, with less invasive procedures, is not enough if not associated with an acceptable level of accuracy. Here, we show the analytical validation behind the Hilab Flow (HiF), a multi-analyte POCT analyzer. HiF quantitative and qualitative tests for 6,175 clinical samples were compared to gold-standard methods from College of American Pathologists accredited laboratories. The compatibility between methods was evaluated in terms of association and clinical agreement. The established approval criteria was a kappa agreement > 0.8. A strong concordance was observed for the 27 analytes tested. Accuracy was greater than 90% for all HiF exams, indicating a good clinical agreement to gold standard laboratory testing. Results indicate that all quantitative and qualitative tests are suitable for POCT and present a reliable performance. HiF stands as a useful tool to aid decision-making in the clinical setting, with potential to contribute to healthcare solutions in diagnostic medicine worldwide.

## 1. INTRODUCTION

Point-of-care testing (POCT) is a form of remote clinical diagnosis with faster turnaround time compared to traditional testing [1–2]. It is a tool evolving as fast as innovation allows, with a potentially transformative impact on healthcare [3]. Current POCT diagnostic devices are equipped with embedded technology and their application in medicine is growing steadily [4]. They optimize clinical decisions, patient outcomes and provide financial advantages [5–6].

POCT offers an alternative to conventional laboratory testing, especially in limited infrastructure settings [7]. Benefits of this model include the use of portable equipment, specimen collection at the test site, immediate identification of biological samples, low sample volume need and fast results, features which can minimize pre-analytical errors and risks related to transport and identification of biological samples [1–2]. For minor conditions and highly prevalent diseases, POCT may be sufficient for medical decisions in the clinical setting, without additional conventional laboratory testing [8].

One of the commercially available POCT devices is the Hilab Flow (HiF), a patented analyzer [9–13] dedicated to interpreting results from lateral flow assays and colorimetric results from vertical flow assays. The small handheld device combines artificial intelligence (AI), machine- and deep-learning techniques to shorten turnaround time, without compromising the test’s performance, the main drawback in general POCT [14]. The system’s flexibility is another important feature of the equipment, which enables performing a variety of laboratory tests.

The main concern regarding POCT is to guarantee reliable results, equivalent to those acquired with standard laboratory-based methods, and under international clinical guidelines [15–17]. Rapid test suppliers usually offer limited information about test performance, however, robust analysis is crucial to offer a high quality product. Therefore, this study aims to provide data from the HiF system analytical validation to assess its performance for POCT use and its potential to aid medical decision-making.

## 2. MATERIALS AND METHODS

### 2.1 Sampling and Data

Internal data from the routine analysis of clinical laboratory service were used for this in-house validation study. Evaluation of respiratory viruses tests, such as COVID-19 Ag, were conducted with nasopharyngeal samples, while other exams were evaluated from whole blood and/or serum specimens. Systematically, except for respiratory viruses analyses, both capillary blood and conventional venous samples were collected from the individuals, at the same time, to the extent of comparing the results of the tests performed on the Hilab Flow reader (capillary) with those from the College of American Pathologists accredited laboratories (venous). The study was approved by the Beneficência Portuguesa Research Ethics Committee: CAEE 33490420.9.0000.5483.

The clinical samples were evaluated by vertical or lateral flow assays, depending on the test type. On vertical flow assays, the presence of the analyte of interest promotes a measurable color change on the test membrane, generated by a chemical reaction between the sample and the test components, which is proportional to the analyte concentration.

Lateral flow assays are membrane-based tests that combine colloidal gold-labeled particles to detect the analyte of interest from nasopharyngeal secretion, whole blood, serum, or plasma specimens. Analyte’s molecules react and form a complex with labeled antibodies or antigens and, as this conjugate moves through capillarity across the membrane, the anti-analyte antibody or antigen immobilized in the membrane binds to the complex, revealing a colored line of varying intensity, that can be measured by its optical density. For qualitative lateral flow tests, a colored test line, accompanied by a control line, indicates a positive result (reagent), while for quantitative tests, the intensity of the test line relates to the analyte’s concentration.

A list of all comparison methods used for clinical correlation is shown in Table 1.

**Table 1.** Analyte, Sample Type, and Methods for Evaluating Clinical Correlation.

| Analyte | HiF |  | Gold Standard |  |  |
| --- | --- | --- | --- | --- | --- |
|  | Assay | Specimen | Assay | Specimen | Equipment |
| COVID-19 IgG/IgM | Lateral Flow | Capillary blood | Chemiluminescence | Serum | Alinity, Abbott |
| COVID-19 Ag | Lateral Flow | Nasopharynx swab | RT-PCR* | Nasopharynx swab | QuantStudio 5, ThermoFisher |
| Influenza A/B Virus Ag | Lateral Flow | Nasopharynx swab | RT-PCR | Nasopharynx swab | QuantStudio 5, ThermoFisher |
| Syphilis Ab | Lateral Flow | Capillary blood | Chemiluminescence | Serum | Alinity, Abbott |
| HIV Ab | Lateral Flow | Capillary blood | Chemiluminescence | Serum | Alinity, Abbott |
| HCV Ab | Lateral Flow | Capillary blood | Chemiluminescence | Serum | Alinity, Abbott |
| HBsAg | Lateral Flow | Capillary blood | Chemiluminescence | Serum | Alinity, Abbott |
| Anti-HBsAg Ab | Lateral Flow | Capillary blood | Chemiluminescence | Serum | Alinity, Abbott |
| Dengue Virus NS1 Ag | Lateral Flow | Capillary blood | Immunochromatography | Serum | Test Strip, Wama |
| Dengue Virus IgG/IgM | Lateral Flow | Capillary blood | Enzyme immunoassay | Serum | Sprinter, Euroimunn |
| Zika Virus IgG/IgM | Lateral Flow | Capillary blood | Enzyme immunoassay | Serum | Sprinter, Euroimunn |
| PSA | Lateral Flow | Capillary blood | Chemiluminescence | Serum | DXI 800, Beckman |
| TSH | Lateral Flow | Capillary blood | Chemiluminescence | Serum | DXI 800, Beckman |
| Beta-hCG | Lateral Flow | Capillary blood | Chemiluminescence | Serum | DXI 800, Beckman |
| 25-Hydroxy Vitamin D | Lateral Flow | Capillary blood | Chemiluminescence | Serum | DXI 800, Beckman |
| HbA1c | Lateral Flow | Capillary blood | HPLC** | Whole blood | Variant, Beckman |
| Glucose | Vertical Flow | Capillary blood | Colorimetric | Serum | AU5800, Beckman |
| Lipid Panel*** | Vertical Flow | Capillary blood | Colorimetric | Serum | AU5800, Beckman |
| Renal Function**** | Vertical Flow | Capillary blood | Colorimetric | Serum | AU5800, Beckman |

\*RT-PCR: Reverse transcription polymerase chain reaction.
\*\*HPLC: High-performance liquid chromatography.
\*\*\*Lipid Panel: cholesterol, HDL-cholesterol, and triglycerides.
\*\*\*\* Renal Function: uric acid, creatinine, and urea.

### 2.2 HiF analyzer

The Hilab Flow (HiF) reader (Hilab, Curitiba-PR) is a laboratory analyzer platform for professional POCT, used for detection and/or quantification of various analytes. The system processes immunochromatography results from lateral flow assays and colorimetric results from vertical flow assays by measuring optical density or color model values, respectively. The equipment is composed of two main parts: a portable handheld analyzer (12 cm x 12 cm x 13 cm, 450 g), which incorporates a camera-equipped light detector, and sample integrated capsules/cartridges. The device communicates with the laboratory’s server, where AI tools and clinical specialists analyze the results. The system applies computer vision and image processing tools to find regions of interest, improve image quality and detect objects. Moreover, machine learning and deep learning techniques perform classification and regression tasks to assist the analysis of quantitative and qualitative exams, respectively. On the field, the current method employs a dual verification of the test result (as explained briefly in the following section). For the sake of evaluating HiF’s accuracy and clinical agreement, all results from this study were based on the robustness of the calibration curves for 27 different analytes. Precision and interference studies are not explored here.

### 2.3 Operation

For HiF testing, a sample (5 µL to 80 µL) is introduced into the capsule by using a capillary tube, pipette, or a medicine dropper device, with an adjuvant buffer (differences apply depending on the exam). The capsule, identified by a unique QR code, is then inserted into the analyzer, which measures the compatible signal of the lateral/vertical flow test. Internet of things technology is used to recognize the QR code from each sample and sends the information regarding the reaction via cloud to the main laboratory. There, an AI software analyses the data, and a licensed health professional, which has the final decision, evaluates and signs the test report. The result is released through the cloud system to the health care service location where the test was performed and via email and/or text message to the patient. Data management and protection are ensured by the system, enabling the tracking of processed samples [12]. The full process occurs within 30 min.

### 2.4 Clinical Agreement Evaluation

To ensure a thorough analysis, each analyte was evaluated in terms of clinical agreement and compatibility between methods. Agreement between quantitative methods was assessed through regression analysis of the plotted curves.

### 2.5 Performance Evaluation

For the HiF performance evaluation, measurements from 10 quantitative and 13 qualitative tests were compared to those from a College of American Pathologists (CAP) accredited laboratory. Some tests are multi-analyte, such as renal function (uric acid, creatinine, urea) and lipid profile (total cholesterol, HDL-c, triglycerides), comprising a total of 27 evaluated analytes. Data regarding detailed analytes, specimens, gold standard assays, and equipment used for comparison and validation of the POCT assays are described in Table 1.

Quantitative clinical correlation analyses were performed for the following analytes: blood glucose, glycosylated hemoglobin (HbA1c), total cholesterol, high-density lipoprotein cholesterol (HDL-c), triglycerides, 25-Hydroxy (25-OH) Vitamin D, thyroid-stimulating hormone (TSH), uric acid, creatinine, and urea. Other analytes from the lipid panel are indirectly estimated through the difference between CHOL and HDL-c (NHDL-c), Martin’s equation (LDL-c), and Friedewald equation (VLDL), but not reported here, because any interpretation on correlation and analytical accuracy would be derived from the relationship among Total Cholesterol, HDL-c, and Triglycerides.

Qualitative clinical correlation analyses were performed for COVID-19 IgG / IgM, COVID-19 antigen (Ag), Influenza A and/or B virus Ag, Syphilis antibody (Ab), human immunodeficiency virus (HIV) Ab, hepatitis C virus (HCV), surface antigen (HBsAg), hepatitis B virus surface Ab (Anti-HBsAg), Dengue virus NS1 Ag, Dengue virus IgG and/or IgM Ab, Zika virus IgG and/or IgM Ab, Beta-human chorionic gonadotropin (Beta-hCG), and prostate-specific antigen (PSA).

### 2.6 Statistical Analysis

The analytical performance and observational errors of the HiF tests were evaluated in terms of clinical agreement parameters, such as accuracy, sensitivity, specificity, and kappa. Analyte total error and regression analysis were evaluated only for the quantitative exams.

The allowable total error evaluation was based on the Clinical Laboratory Improvement Amendments as indicated in Equation 1 (CLIA, 2019). Predicted and measured values refer to the analyte concentration reported for the measured (gold standard assay) and proposed method (HiF Method), respectively.

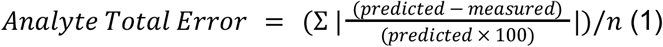

The intercept and the slope coefficients and errors of the regression were evaluated using a linear model and the correlation coefficient *r* was obtained from a spearman correlation test, since data showed to be non-parametric according to Shapiro-Wilk tests and Q-Q plot analysis.

The ranges for the clinical agreement analysis were defined, as shown in Table 2. Intermediate values of clinical interpretation for TSH (primary and mild hypothyroidism), HbA1c (pre-diabetes), and Glucose (pre-diabetes and diabetes) exams were included in the same category to provide binary outcomes and simplify the analysis.

**Table 2.** Interpretation and Clinical Ranges for Clinical Agreement Assessment.

| Test / Analyte |  | Interpretation / Clinical Ranges |  |  | Measuring Range |
| --- | --- | --- | --- | --- | --- |
| COVID-19 IgG/ IgM | Reactive | Non-reactive | - | - | N/A |
| Syphilis | Reactive | Non-reactive | - | - | N/A |
| Anti-HBsAg | Reactive | Non-reactive | - | - | N/A |
| Zika IgG / IgM | Reactive | Non-reactive | - | - | N/A |
| Influenza A / B | Reactive | Non-reactive | - | - | N/A |
| HIV | Reactive | Non-reactive | - | - | N/A |
| HCV | Reactive | Non-reactive | - | - | N/A |
| HBsAg | Reactive | Non-reactive | - | - | N/A |
| Dengue NS1 | Reactive | Non-reactive | - | - | N/A |
| COVID-19 Ag | Reactive | Non-reactive | - | - | N/A |
| Dengue IgG/ IgM | Reactive | Non-reactive | - | - | N/A |
| PSA | Reactive | Non-reactive | - | - | N/A |
| Beta-hCG | Pregnancy | Non-Pregnancy | - | - | N/A |
| Total Cholesterol (mg/dL) | High (> 190 mg/dL) | Normal (< 190) | - | - | 100 - 400 |
| HDL-c (mg/dL) | Low (< 40) | Normal (> 40) | - | - | 20 - 100 |
| Triglycerides (mg/dL) | High (> 150) | Normal (< 150) | - | - | 100 - 400 |
| HbA1c (%) | Diabetes (> 6.5)** | Pre-Diabetes (5.7-6.4)** | Normal (< 5.7) | - | 4 - 10 |
| Glucose (mg/dL) | Diabetes (> 126 mg/dL) | Pre-Diabetes (100-126)** | Normal (70-100)** | Hypoglycemia (< 70)** | 20 - 400 |
| 25-OH Vitamin D (ng/mL) | Deficiency (< 20) | Normal (20-60) | - | - | 10 - 70 |
| TSH (μIU/mL) | Primary Hypothyroidism (> 10)** | Mild Hypothyroidism (4.5-10)** | Normal (< 4.5) | - | 4.5 - 40 |
| Uric Acid (mg/dL) | High (male > 7<br>female > 5.7) | Normal (male < 7<br>female < 5.7) | - | - | 3.5 - 12 |
| Creatinine (mg/dL) | High (male > 1.3<br>female > 1.1) | Normal (male < 1.3<br>female < 1.1) | - | - | 0.6 - 4 |
| Urea (mg/dL) | High (> 43) | Normal (< 43) | - | - | 10 - 100 |

\* Considered in the same category

The analysis was conducted in R (R Core Team, 2022) Significance level was fixed at 0.05. The dataset is available under request. Please, contact.

## 3. RESULTS AND DISCUSSION

Universal healthcare availability depends on decentralized diagnostic POCT systems both in wealthy and developing countries [20–21]. Different from their predecessors, modern POCT devices are smaller, smarter, easier to use, less prone to errors, and go without additional testing for clinical purposes [4]. However, this scenario requires a reliable and robust clinical agreement with an established laboratory method [22–23]. Thus, combining robust analytical validation and reliable quality control in the clinical laboratory is the key to improving healthcare access.

To ensure a thorough analysis, each test was evaluated in terms of analytical performance, analytical sensitivity, and compatibility between methods. Most analyses showed total error within the parameters suggested by CLIA’s guidelines on analytical performance. For the remaining analytes (Glucose and Renal Function analytes), no systematic difference was observed between HiF and conventional methods after applying the appropriate statistical test and evaluating their clinical performance (Table 3).

**Table 3.** Comparison of HiF Total Error (%) and Allowable Total Error (CLIA) in Terms of Analyte Biological Variation for Quantitative Assays.

| Analyte | Analyte Total Error (%) | Allowable Total Error (%) |
| --- | --- | --- |
| Total Cholesterol | 9.35% | 10.0% |
| HDL-c | 12.01% | 20.0% |
| Triglycerides | 7.13% | 15.0% |
| HbA1c | 10.19% | 10.0% |
| Glucose | 9.92% | 8.0% |
| 25-Hydroxy Vitamin D* | 21.96% | 25.0% |
| TSH | 3.74% | 20.0% |
| Uric Acid | 20.36% | 10.0% |
| Creatinine | 19.49% | 10.0% |
| Urea | 16.87% | 9.0% |

\* The allowable total error for 25-hydroxy Vitamin D is not available in CLIA Guidelines, thus, it was based on Stöckl, Sluss & Thienpont (2009).

Due to a lack of consensus in literature, we followed IUPAC and CLSI definitions for “analytical sensitivity”, which refers to the concept of assay sensitivity as slight changes in analyte concentration [24]. That is, the closer to 1 is the slope of the regression curve, the more precise the method. Several guidelines on method validation highlight the importance of calculating regression parameters (e.g. correlation coefficient, y-intercept) for a comprehensive method validation [25–26], but no clear criteria are established for the acceptable range of estimated slope, especially for POCT [27]. Thus, our acceptance criteria for assay method agreement was obtaining a slope anywhere between 0.85 - 1.15, which is a common rule of thumb for most rapid test strip suppliers (Figure 1).

**Figure 1.**
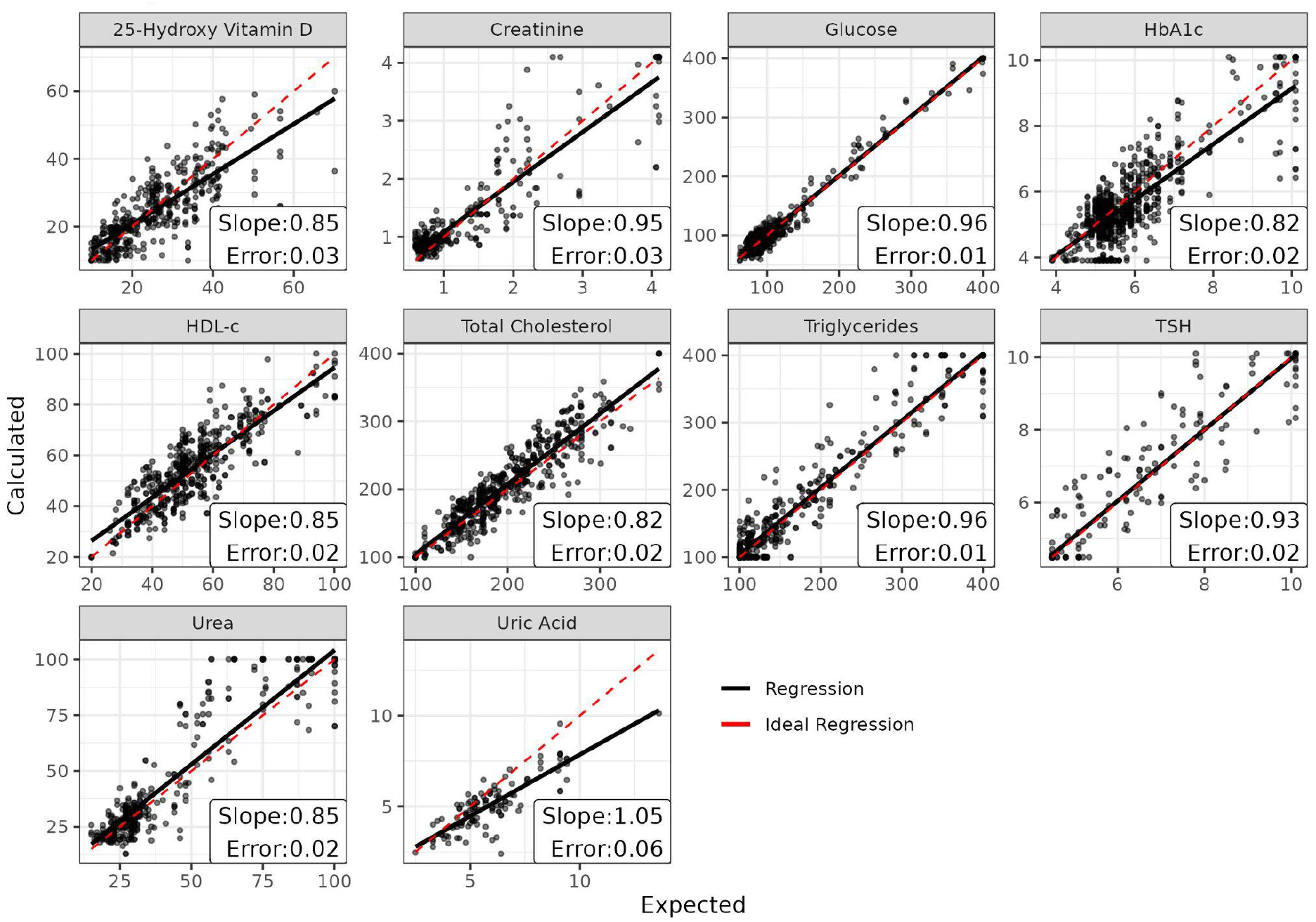
Linear Regression Analysis, slope coefficient, and error from the calculated value. All tests matched our criteria since estimated slopes (with confidence intervals) ranged from 0.85 to 1.15. For HbA1c and Total Cholesterol, a greater variation in slope did not impact the clinical agreement regarding decision limits, as indicated by resulting sensitivity and specificity, described next.

POCT is a unique methodology, with different biases and limitations when compared to conventional laboratory testing [29–30]. We considered an acceptable variation (between Hilab and cut-off value) for results within 5% of a category boundary to avoid overestimation of clinical differences [28]. CLIA [31] acceptable analytical performance greatly varies depending on the evaluated analyte (5 - 30%), but, we decided on 5%, regardless of the analyte, to enable a more accurate (and less biased) performance estimation. In this context, most analytes showed performance parameters greater than 80% (sensitivity, specificity, and accuracy), suggesting a good agreement for the HiF method (Tables 4 and 5). In the case of Uric Acid, the low number of altered samples (20% in total) and unbalanced proportion (altered VS normal) probably impacted on estimated Kappa and sensitivity.

**Table 4.** Clinical Agreement for Quantitative Assays.

| Analyte | Sensitivity | Specificity | Accuracy | Kappa | n |
| --- | --- | --- | --- | --- | --- |
| **TSH** | 1.000 (0.995-1.000) | 0.991 (0.986-0.997) | 0.994 (0.980-0.999) | 0.990 | 574 |
| **Total Cholesterol** | 0.992 (0.989-0.996) | 0.922 (0.919-0.925) | 0.957 (0.935-0.972) | 0.910 | 529 |
| **HDL-c** | 0.670 (0.668-0.672) | 0.955 (0.951-0.959) | 0.900 (0.870-0.925) | 0.660 | 499 |
| **Triglycerides** | 0.980 (0.977-0.984) | 0.954 (0.951-0.957) | 0.963 (0.944-0.977) | 0.920 | 570 |
| **25-Hydroxy Vitamin D** | 0.884 (0.879-0.888) | 0.938 (0.933-0.942) | 0.918 (0.887-0.943) | 0.820 | 402 |
| **HbA1c** | 0.818 (0.816-0.820) | 0.972 (0.970-0.975) | 0.947 (0.930-0.962) | 0.810 | 819 |
| **Glucose** | 0.928 (0.923-0.932) | 0.997 (0.993-1.000) | 0.986 (0.970-0.995) | 0.950 | 428 |
| **Uric Acid** | 0.636 (0.627-0.645) | 1.000 (0.983-1.000) | 0.930 (0.866-0.969) | 0.740 | 114 |
| **Creatinine** | 0.926 (0.919-0.932) | 0.942 (0.936-0.949) | 0.935 (0.899-0.961) | 0.870 | 277 |
| **Urea** | 0.936 (0.931-0.941) | 0.981 (0.975-0.986) | 0.961 (0.936-0.979) | 0.920 | 363 |
|  |  |  |  |  | **4575** |

**Table 5.** Clinical Agreement for Qualitative Assays.

| Analyte | Sensitivity | Specificity | Accuracy | Kappa | n |
| --- | --- | --- | --- | --- | --- |
| **COVID IgG/ IgM** | 1.000 (0.990-1.000) | 1.000 (0.990-1.000) | 1.000 (0.982-1.000) | 1.000 | 200 |
| **Syphilis** | 1.000 (0.980-1.000) | 1.000 (0.980-1.000) | 1.000 (0.964-1.000) | 1.000 | 100 |
| **Anti-HbsAg** | 0.964 (0.945-0.983) | 1.000 (0.980-1.000) | 0.980 (0.930-0.998) | 0.960 | 100 |
| **HIV** | 1.000 (0.980-1.000) | 1.000 (0.980-1.000) | 1.000 (0.964-1.000) | 1.000 | 100 |
| **HCV** | 1.000 (0.980-1.000) | 1.000 (0.980-1.000) | 1.000 (0.964-1.000) | 1.000 | 100 |
| **HBsAg** | 1.000 (0.980-1.000) | 1.000 (0.980-1.000) | 1.000 (0.964-1.000) | 1.000 | 100 |
| **PSA** | 0.897 (0.879-0.914) | 1.000 (0.980-1.000) | 0.940 (0.874-0.978) | 0.880 | 100 |
| **COVID-19 Ag** | 1.000 (0.980-1.000) | 1.000 (0.980-1.000) | 1.000 (0.964-1.000) | 1.000 | 100 |
| **Dengue NS1** | 1.000 (0.990-1.000) | 1.000 (0.990-1.000) | 1.000 (0.982-1.000) | 1.000 | 200 |
| **Influenza A/ B** | 1.000 (0.990-1.000) | 1.000 (0.990-1.000) | 1.000 (0.982-1.000) | 1.000 | 200 |
| **Dengue IgG/ IgM** | 1.000 (0.990-1.000) | 1.000 (0.990-1.000) | 1.000 (0.982-1.000) | 1.000 | 200 |
| **b-HCG** | 1.000 (0.980-1.000) | 0.985 (0.966-1.000) | 0.990 (0.946-1.000) | 0.980 | 100 |
|  |  |  |  |  | **1600** |

**Table 6.** Summary of Analytical Performance and Clinical Agreement Between Methods.

| **Analyte** | **Kappa** | **Analytical Performance** |
| --- | --- | --- |
| **TSH** | > 0.9 | Within CLIA Limits |
| **Total Cholesterol** | > 0.9 | Within CLIA Limits |
| **HDL-c** | < 0.8 | Within CLIA Limits |
| **Triglycerides** | > 0.9 | Within CLIA Limits |
| **25-Hydroxy Vitamin D** | > 0.8 | Within CLIA Limits |
| **HbA1c** | > 0.8 | 0.2% Off CLIA Limits |
| **Glucose** | > 0.9 | 2% Off CLIA Limits |
| **Uric Acid** | < 0.8 | 10% Off CLIA Limits |
| **Creatinine** | < 0.8 | 10% Off CLIA Limits |
| **Urea** | > 0.8 | 8% Off CLIA Limits |
| **Qualitative Exams** | > 0.8 | N/A* |

\* The following parameter is only eligible for quantitative assays.

In summary, the HiF shows adequate clinical agreement for all qualitative exams, expressed in terms of sensitivity and specificity. For the quantitative exams, all analytes (but HDL-c and Uric Acid) achieved a good method agreement (Kappa > 0.8) and analytical performance within CLIA acceptable variation. Estimated total error (%) for HbA1c and Glucose was slightly higher than the CLIA Limits (0.2% and 2%), respectively, but, without real impact on exams sensitivity and specificity. Finally, all renal function analytes (Uric Acid, Creatinine, and Urea) presented only a moderate clinical agreement or analytical performance, suggesting their use for screening purposes only.

This study has limitations. First, we focus on reporting clinical correlation data rather than approaching precision, interference, and limit of detection/ quantification studies. Although these are important steps in method validation, we chose not to approach them here. Instead, we highlight the clinical agreement of the Hilab assays, precisely, the ultimate motivation of this study. Another potential limitation is a result bias due to the use of different types of samples. For most analyzes, the HiF tested for whole blood, while for validation in a reference laboratory, the standard sample was serum. Complementary studies have been carried out to determine the performance of the HiF reader about these variables, but are not represented here since the sample size would prevent a relevant statistical analysis.

In the POCT industry, devices are usually designed to analyze a small set of analytes, which limits the technology’s spreading. The HiF analyzer enables a multi-analyte evaluation on a single device for quantitative and qualitative laboratory tests. Also, the system employs a dual verification of results (by AI and a licensed specialist), minimizing common analytical errors from decentralized laboratories. Previous works from our group show AI can improve the analytical precision of a method, thus enabling epidemiological conclusions on a populational level [13, 32–33]. The effective combination of internet of things and AI tools for POCT offers, in fact, an alternative to conventional laboratory testing [34].

## 4. CONCLUSION

This study presents data from 6,175 clinical samples used to evaluate the clinical agreement between a POCT-based analyzer (HiF) and a CAP-certified laboratory. The HiF reader combines internet of things and AI tools to achieve reliable performance, representing an alternative to conventional laboratory testing. In this study, a thorough evaluation, comprising regression analysis and clinical agreement, revealed the performance characteristics of the device, to ensure the patient gets the best outcome from the technology. Results show that the HiF system is useful to aid medical decision-making in the clinical setting, with the potential to contribute to healthcare solutions in diagnostic medicine.

## Data Availability

All data produced in the present work are contained in the manuscript.

## CRediT author statement

**Lucca Centa Malucelli:** Project Administration, Writing, Editing, and Review (Original and Final draft).

**Gabriele Luise Neves Alves:** Writing, Editing, and Review (Original and Final draft).

**Claucio Antonio Rank Filho:** Writing, Editing, and Review (Original and Final draft).

**Rafaela Fortes Correa:** Laboratory analysis and testing.

**Vanessa Hintz Albano:** Laboratory analysis and testing.

**Carolina Melchioretto dos Santos:** Supervision.

**Matheus Gonçalves Severo:** Editing and Review (Original and Final draft).

**Victor Henrique Alves Ribeiro:** Writing, Editing, and Review (Final draft).

**Bernardo Montesanti Machado de Almeida:** Writing, Editing, and Review (Original and Final draft), Supervision.

**Caio Corsi Klosovski:** Editing and Review (Original and Final draft).

**Tania Leme da Rocha Martinez:** Editing and Review (Original and Final draft), Supervision.

**Marileia Scartezini:** Editing and Review (Original and Final draft), Supervision.

**Tereza Luiza Bellincanta Fakhouri:** Editing and Review (Original and Final draft).

**Marcus Vinícius Mazega Figueredo:** Funding Acquisition and Supervision.

## ACKNOWLEDGMENT

We acknowledge funding for the analytical method validation of the tests reported here.

## REFERENCES

[1] Larsson, A., Greig-Pylypczuk, R., Huisman, A. The state of point-of-care testing: a european perspective, Upsala Journal of Medical Sciences. 2015, 120(1): 1–10.

[2] Luppa, PB., Müller, C., Schlichtiger, A., Schlebusch, H. Point-of-care testing (POCT): Current techniques and future perspectives. Trends in Analytical Chemistry. 2011, 30(6): 887–898.

[3] Jani, IV., Peter, TF. How point-of-care testing could drive innovation in global health. New England Journal of Medicine. 2013, 368(24): 2319–24.

[4] St John, A., Price, CP. Existing and Emerging Technologies for Point-of-Care Testing. Clin Biochem Rev. 2014, 35(3): 155–67.

[5] St John, A., Price, CP. Economic evidence and point-of care testing. Clin Biochem Rev. 2013, 34:61–74.

[6] Laurence, CO., Moss, JR., Briggs, NE., Beilby, JJ., Po, CT. PoCT Trial Management Group. The cost-effectiveness of point of care testing in a general practice setting: results from a randomised controlled trial. BMC Health Serv Res. 2010, 10: 165.

[7] Kozel, TR., Burnham-Marusich, AR. Point-of-Care Testing for Infectious Diseases: Past, Present, and Future. J Clin Microbiol. 2017, 55(8): 2313–2320.

[8] Park, HD. Current Status of Clinical Application of Point-of-Care Testing. Arch Pathol Lab Med. 2021, 145(2): 168–175.

[9] Figueredo, M. V. M., Junior, S. R. R., Cossetin, M. J., Cemin, R. S. C., Pinto, R. N., Santos, A. R., Andrade, G. H. B., Camati, M. E. M. (2018). Leitor de Amostras Biológicas em Testes Bioquímicos Rápidos. BR 102017008428-0 A2. instituto Nacional da Propriedade Industrial.

[10] Figueredo, M. V. M., Junior, S. R. R., Cossetin, M. J., Cemin, R. S. C., Pinto, R. N., Santos, A. R., Andrade, G. H. B., Camati, M. E. M. (2018). Sistema de Coleta, Exame e Leitura de Amostras Biológicas em Testes Bioquímicos Rápidos e Gerenciamento de Dados. Instituto Nacional da Propriedade Industrial.

[11] Figueredo, M. V. M., Junior, S. R. R., Cossetin, M. J., Cemin, R. S. C., Pinto, R. N., Santos, A. R., Andrade, G. H. B., Camati, M. E. M. (2018). Reader Device for Biological Samples in Rapid Biochemical Tests. WO 2018/195616 Al. Intellectual Property Office.

[12] Figueredo, M. V. M., Junior, S. R. R., Cossetin, M. J., Cemin, R. S. C., Pinto, R. N., Santos, A. R., Andrade, G. H. B., Camati, M. E. M. (2020). Biological Sample Collecting, Examining and Reading System for Fast Biochemical Assays and Data Management. System for biological sample collection, examination and reading in quick biochemical tests and data management. WO 2018/195617 Al. Intellectual Property Office.

[13] Cardoso, C. Q., Almeida. B. M. M., Saldanha, A. L. R., Klosovski, C. C., Margeotto A. P. P., Gasparoto, A. L. V., Scartezini, M., Martinez, T. L. R. (2021). Brazilian Lipid Cardiovascular Risk Pre and During the Covid 19 Pandemic in Asymptomatic and Severely Affected Groups. Online Journal of Cardiology Research & Reports. DOI: 10.33552/OJCR.2021.05.000606.

[14] Florkowski, C., Don-Wauchope, A., Gimenez, N., Rodriguez-Capote, K., Wils, J., & Zemlin, A. (2017). Point-of-care testing (POCT) and evidence-based laboratory medicine (EBLM)–does it leverage any advantage in clinical decision making?. Critical reviews in clinical laboratory sciences, 54(7-8), 471–494.

[15] Pecoraro, V., Banfi, G., Germagnoli, L., Trenti, T. A systematic evaluation of immunoassay point-of-care testing to define impact on patients’ outcomes. Ann Clin Biochem. 2017, 54(4): 420–431.

[16] International Organization for Standardization (ISO) 22870:2006 Point-of-care testing care (POCT) – Requirements for quality and competence. Geneva: ISO, 2006.

[17] CLSI. Quality Management: Approaches to Reducing Errors at the Point of Care. 1st Edition. CLSI guideline POCT07. Wayne, PA: Clinical and Laboratory Standards Institute, 2010.

[18] Agresti, A., Coull, BA. Approximate is better than “exact” for interval estimation of binomial proportions. The American Statistician. 1998, 52(2): 119–126.

[19] Watson, PF., Petrie, A. Method agreement analysis: a review of correct methodology. Theriogenology. 2010, 73(9): 1167–1179.

[20] Urdea, M., Penny, LA., Olmsted, SS., Giovanni, MY., Kaspar, P., Shepherd, A., Wilson, P., Dahl, CA., Buchsbaum, S., Moeller, G., Hay Burgess, DC. Requirements for high impact diagnostics in the developing world. Nature. 2006, 444(1): S73–S79.

[21] Pai, NP., Vadnais, C., Denkinger, C., Engel, N., Pai, M. Point-of care testing for infectious diseases: diversity, complexity, and barriers in low- and middle-income countries. PLoS Med. 2012, 9: e1001306.

[22] Price, C., St. John, A., Kricka, L. Point-of-Care Testing. Needs, opportunities and innovation. 3rd Edition. Washington, USA: AACC Press, 2010.

[23] Hsieh, YH., Gaydos, CA., Hogan, MT., Uy, OM., Jackman, J., Jett-Goheen, M, et al. What qualities are most important to making a point of care test desirable for clinicians and others offering sexually transmitted infection testing? PLoS One. 2011, 6: e19263.

[24] NCCLS. Protocols for Determination of Limits of Detection and Limits of Quantitation; Approved Guideline. NCCLS document EP17-A (ISBN 1-56238-551-8). NCCLS, 940 West Valley Road, Suite 1400, Wayne, Pennsylvania 19087-1898 USA, 2004.

[25] ICH. International Conference on Harmonization. Guidance for industry: Q2B validation of analytical procedures: methodology. International Council on Harmonization of Technical Requirements for Registration of Pharmaceuticals for Human Use, 1996.

[26] VICH. Validation of analytical procedures: methodology (VICH GL2). 1999.

[27] Krause, Stephan O. Good analytical method validation practice: Deriving acceptance criteria for the AMV protocol: Part II. Journal of validation technology. 2003, 9(2):. 162–179.

[28] Ferreira, CE., França, CN., Correr, CJ., Zucker, ML., Andriolo, A., Scartezini, M. Clinical correlation between a point-of-care testing system and laboratory automation for lipid profile. Clinica Chimica Acta. 2015, 446: 263–266.

[29] Armbruster, D., Miller, R. R. (2007). The Joint Committee for Traceability in Laboratory Medicine (JCTLM): a global approach to promote the standardization of clinical laboratory test results. Clin. Biochem. Rev., 28(3): 105–114.

[30] Wiencek, J., Nichols, J. (2016). Issues in the practical implementation of POCT: overcoming challenges. Expert Review of Molecular Diagnostics, 16(4): 415–422.

[31] US Department of Health and Human Services (HHS). Clinical laboratory improvement amendments of 1988 (CLIA) proficiency testing regulations related to analytes and acceptable performance, 2019.

[32] Martinez, T. L. R., Almeida, B. M. M., Cardoso, C. Q., Saldanha, A. L. R., Scartezini, M., Klosovski, C. C., Pereira, A., Santos Filho, R. D. (2021). The importance of the ratio triglycerides/HDL-c C in RHE Brazilian population residual risk. Atherosclerosis, 331, e204.

[33] Ribeiro, V. H. A., Steinhaus, G., Severo, E. B., Junior, J. R. F., Barbosa, L. J. L., Cossetin, M., & Figueiredo, M. V. M. (2021). A System for Enhancing Human-level Performance in COVID-19 Antibody Detection. In Anais do XXI Simpósio Brasileiro de Computação Aplicada à Saúde (pp. 224–233). SBC.

[34] Gasparin, A. T., Araujo, C. I. F., Schmitt, P., Cardoso, M. R., Perussolo, M. C., de Jesus, T. C. S., Santiago, E. B., Silva, I. L. R., de Sousa, R. G., Teng, F. Z., Severo, E. B., Ribeiro, V. H. A., Cardoso, M. A., Silva, F. D., Perazzoli, C. R. A., Farias, J. S. H., Almeida, B. M. M., Júnior, S. R. R, Figueredo, M. V. M. (2022). Hilab system, a new point-of-care hematology analyzer supported by the Internet of Things and Artificial Intelligence. Scientific reports, 12(1), 1–11.

